# The prevention of nosocomial SARS-CoV2 transmission in endoscopy: a systematic review of recommendations within gastroenterology to identify best practice

**DOI:** 10.1101/2020.03.17.20037564

**Authors:** John Ong, Gail Brenda Cross, Yock Young Dan

**Affiliations:** Department of Medicine, Yong Loo Lin School of Medicine, National University of Singapore, Kent Ridge Road, Singapore; Department of Engineering (Materials Engineering & Materials-Tissue Interactions Group), University of Cambridge, Trumpington Street, Cambridge, UK; East of England Deanery Gastroenterology Training Programme, Cambridge, UK; Division of Infectious Diseases, University Medicine Cluster, National University Hospital, Lower Kent Ridge Road, Singapore

## Abstract

Endoscopy generates aerosol droplets and fomites, thereby increasing the risk of SARS-CoV2 transmission to healthcare workers and uninfected patients within endoscopy departments. Despite the sharp rise in the incidence of COVID-19, authoritative recommendations to limit the spread of SARS-CoV2 within gastrointestinal endoscopy units are lacking. Therefore, with the primary aim of identifying best practice and scrutinizing its supporting evidence, we conducted a systematic review of literature for articles published between 1 January 2002 and 15 March 2020 in five databases relating to both the current SARS-CoV2 and the previous SARS-CoV outbreaks. Official websites for gastroenterology and endoscopy societies in the 15 most affected countries were also searched. Unfortunately, a paucity of high quality data and heterogeneity of recommendations between countries was observed. Interestingly, not all countries advocated the postponement of non-urgent or elective procedures. Recommendations for patient screening and personal protective equipment were commonly featured in all recommendations but specifics varied. Only 32% (9/28) of all gastroenterology and endoscopy societies issued guidance on endoscopy in the COVID-19 pandemic. In conclusion, stronger evidence to inform current practice and robust guidelines are urgently needed to prevent the transmission of SARS-CoV2 in gastrointestinal endoscopy departments worldwide.

## INTRODUCTION

SARS-CoV2 has spread to all major continents. Over 100,000 individuals have been infected and new cases are rising at an alarming rate. Over 3000 healthcare workers (HCW) in China alone are suspected of COVID-19 and over 1,700 tested positive.[1] These statistics underline the imperative need to define appropriate protective guidelines for HCWs in high risk specialties such as gastroenterology to prevent the transmission of SARS-CoV2 both to patients and colleagues.

Transmission of SARS-CoV2 is postulated to be through aerosol droplets or fomites and gastrointestinal endoscopy are high-risk procedures because of aerosolization of bodily secretions. Pharolaryngeal irritation during upper gastrointestinal endoscopy generates aerosol droplets each time a patient coughs or gags. Belching and flatulence caused by insufflation during endoscopy may also generate aerosol droplets. In addition, COVID-19 mimics gastrointestinal disease and infected patients can present unknowingly to endoscopy units if not appropriately screened; up to 20% of COVID-19 patients present with diarrhoeal illness, approximately 5% have nausea and vomiting, and those in the early phases of infection may be asymptomatic but still infectious.[2,3] A single virus-shedding COVID-19 patient with a high viral load can contaminate an entire endoscopy unit including personal protective equipment (PPE), putting HCWs and patients at risk.[4]

Singapore at one point in time had the largest cohort of infected patients outside China in the early phases of the SARS-CoV2 outbreak. Given the novelty of the disease, the quality of preventative measures implemented within our endoscopy units was unknown. Therefore, with the primary aim of identifying best practice in current literature to curb the spread of SARS-CoV2, we performed a systematic review to scrutinize the evidence and practice protocols related to both COVID-19 and Severe Acute Respiratory Syndrome (SARS).

## METHODS

### Search strategy and article selection

A systematic search (Fig.1) using the terms “Severe Acute Respiratory Syndrome”, “SARS”, “SARS-CoV”, “SAR-CoV2”, “COVID-19” and “coronavirus”, in combination with, “endoscopy”, “gastroscopy”, “oesophago-gastro-duodenoscopy”, “esophago-gastro-duodenoscopy”, “sigmoidoscopy”, “colonoscopy”, “ERCP”, “endoscopic retrograde cholangiopancreatography”, and “enteroscopy” were performed in five database (PubMed, Scopus, Cochrane, bioRxiv, and medRxiv) for articles published from 1 January 2002 to 15 March 2020. In the event guidelines were not yet indexed in these databases, official websites for gastroenterology and endoscopy societies (*n* = 28) from the 15 most-affected countries listed by the European Center for Disease Prevention and Control (https://www.ecdc.europa.eu/en) were also searched (Supplementary Table 1).

**Figure 1.**
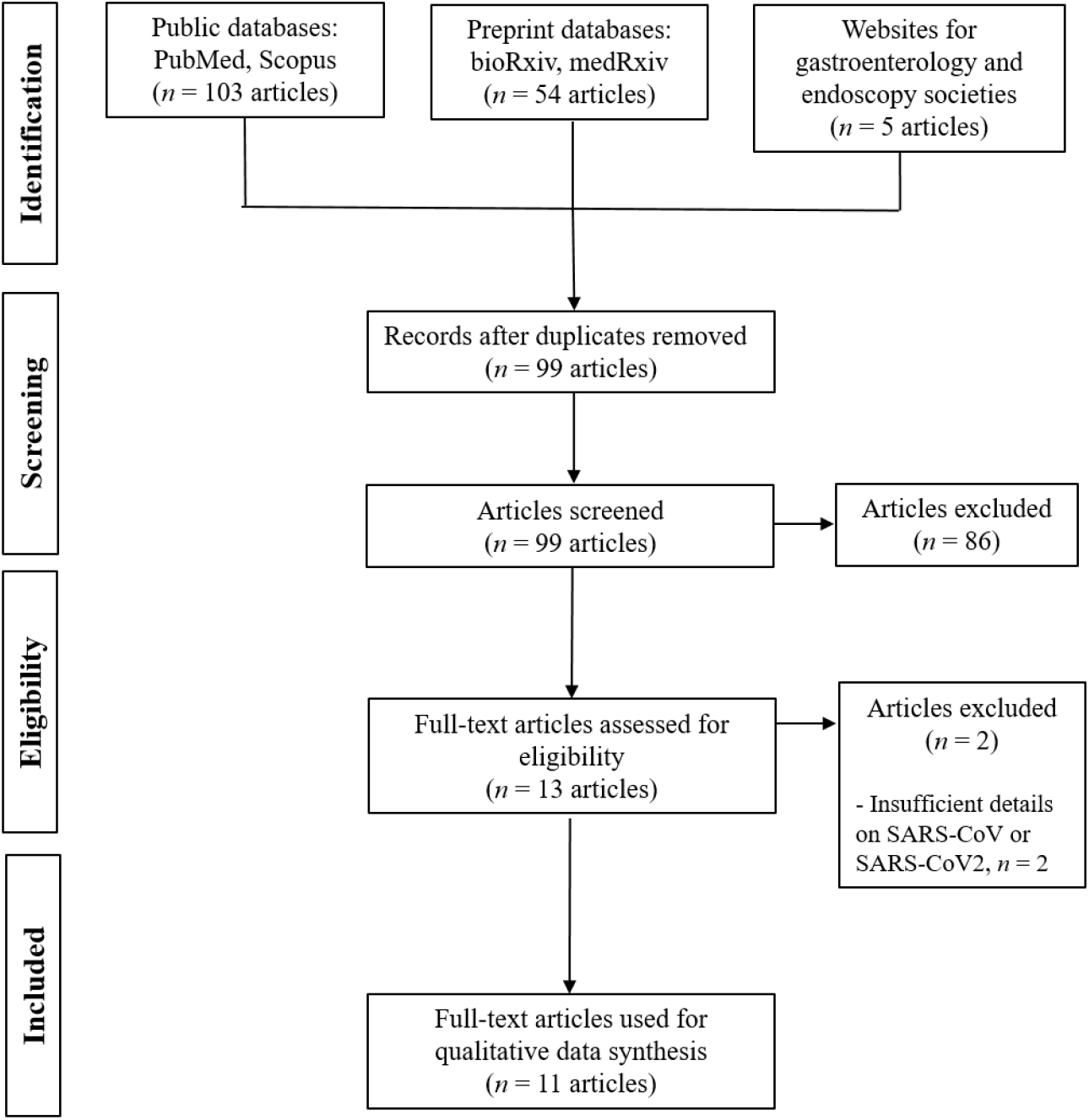
Flow diagram of systematic searches, articles include for review, and artiticles included for qualitative synthesis.

The search was conducted by 2 reviewers independently (JO and YYD). All articles types were screened by abstracts. Duplicates and irrelevant articles were removed. There were no language restrictions. Included articles that were not in English were translated. Extracted data were analyzed for qualitative synthesis by JO and YYD. Any disagreements were resolved by discussions between all three authors.

### Outcome assessment

Our primary outcome was the effect of preventative measures on the incidence of COVID-19 cases within endoscopy departments. Our secondary outcome was the quality of recommendations for (i) patient selection including screening, (ii) peri- and intra-endoscopy practices, and (iii) post-procedure practices.

### Eligibility selection and data extraction

Data from included articles was extracted independently by JO and YYD using pre-designed forms on Microsoft Word (2007 Home Edition; Microsoft Corp, Redmond, Washington). In the event quantitative data was not reported to achieve our primary objective, articles were still scrutinized for data to achieve our secondary objective. Articles with missing data that could not satisfy both primary and secondary objectives were excluded from qualitative synthesis.

### Quality Assessment

The Newcastle-Ottawa Scale was used to assess the quality of the only article that provided quantitative data on the intervention of protective measures.[6] This scored 3 stars for selection, no stars for comparability, and 1 star for outcome, making it a poor-quality study.

### Statistical analysis and qualitative synthesis

There was insufficient data in current literature to perform any statistical analyses to meet our primary objective. For our secondary objective, qualitative analyses involved the stringency and level of detail in the recommendations across three domains: patient selection, peri-procedural and intra-procedural practices, and post-procedural practices. A fourth domain “general advice” was created to report any useful data which did not fit the previous three domains. For patient selection, screening protocols (e.g. temperature readings, imaging, etc.), contingency plans for a high-risk patient newly detected in endoscopy (suspected or positive patient), triaging, and recommendations for PPE (patients and front desk staff) were assessed. For peri-procedural and intra-procedural practices, the recommendation for PPEs and infection control measures were assessed. For post-procedural practices, decontamination practices and recommended PPE for transfer staff. Monitoring of staff and contingency plans for unprotected HCWs post-exposure were assessed and included under “general advice”. All authors contributed to the qualitative synthesis.

## RESULTS

### Search results

9 guidelines [4-12] and 2 articles [13-14] on preventative measures during the SARS outbreak were reviewed for qualitative synthesis. 9 of 10 guidelines related to the COVID-19, the other was the American Society for Gastrointestinal Endoscopy (ASGE) 2003 recommendation for the SARS outbreak [12]. Of the 8 COVID-19 guidelines, 3 originated from China, 2 from US, 2 from UK and 1 from Spain. Of all gastrointestinal-related societies reviewed, 32% (9/28) had published advice on the management of suspected or confirmed COVID-19 cases at the point of writing. Only 1 of all reviewed articles cited the efficacy of its preventative measures on the incidence of COVID-19 cases, however, sample size was small and period of observation abrupt (See Table 1). There was insufficient data in literature for meta-analyses. Breadth of recommendations and depth of detail varied considerably in all domains between countries, being most stringent in China.

**Table 1:**
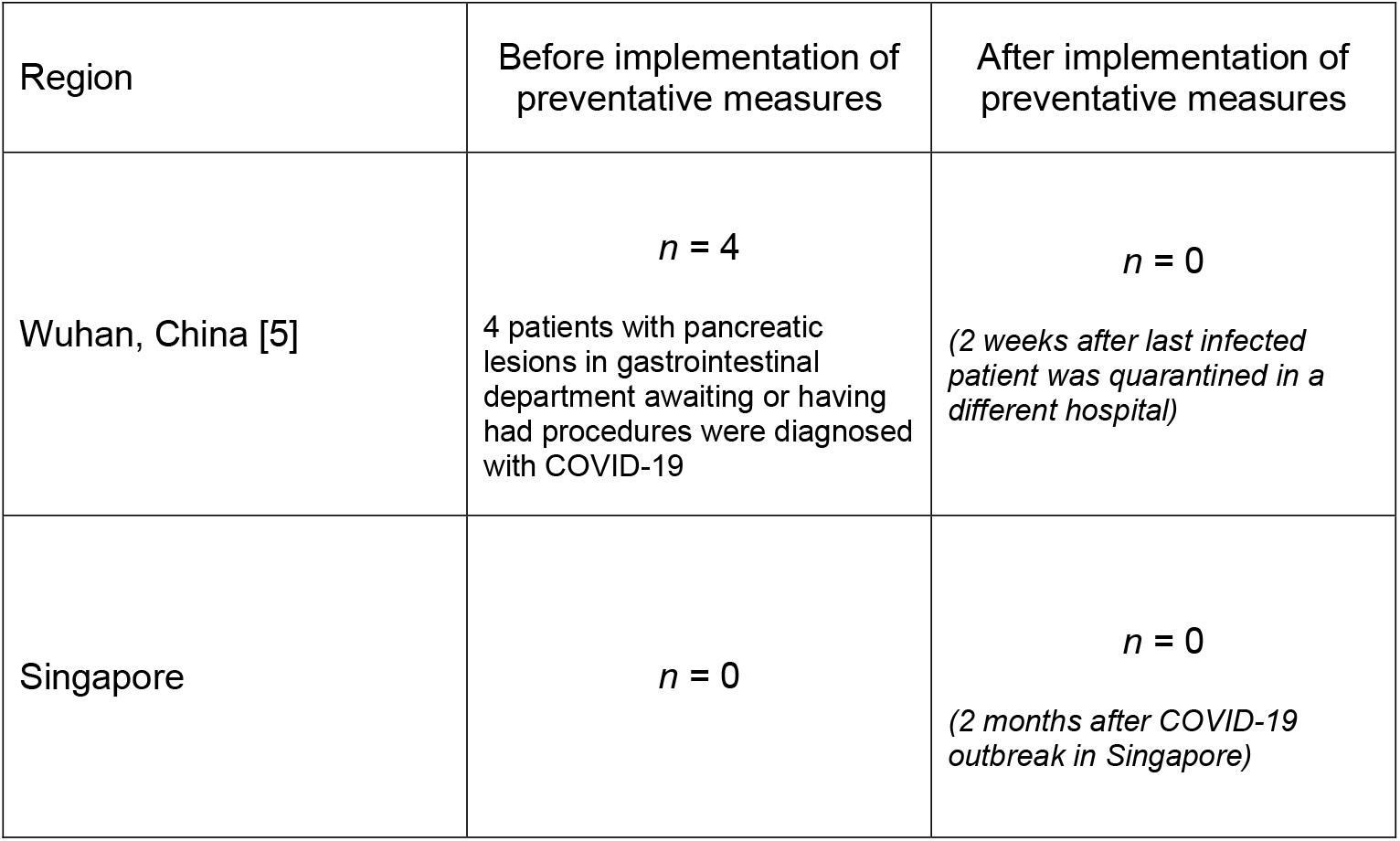
The incidence COVID-19 cases (patients or HCWs) in gastrointestinal or endoscopy departments before and after the implementation of preventative measures – displayed with results from Singapore for comparison.

| Region | Before implementation of preventative measures | After implementation of preventative measures |
| --- | --- | --- |
| Wuhan, China [5] | $n = 4$<br>4 patients with pancreatic lesions in gastrointestinal department awaiting or having had procedures were diagnosed with COVID-19 | $n = 0$<br>*(2 weeks after last infected patient was quarantined in a different hospital)* |
| Singapore | $n = 0$ | $n = 0$<br>*(2 months after COVID-19 outbreak in Singapore)* |

### Patient selection

Details of patient selection for endoscopy has been summarised in Table 2. Not all guidelines and countries recommended the postponement of non-urgent or elective procedures. In Spanish and British guidelines, cases referred to endoscopy were primarily triaged by patient risk of having COVID-19. In contrast to US, China and Singapore, endoscopy cases were firstly triaged by clinical need then by patient risk of COVID-19. In terms of screening protocols, all protocols advocated the use of body temperature > 37.5°C, symptoms of COVID-19, travel history and contact history. Recommendations from China appeared to be most stringent and advocated the use of chest computerised tomography (CT Lung) and real-time polymerase chain reaction (RT-QCR) in suspicious cases. Apart from Chinese related guidelines that recommended the isolation of all positive patients detected through endoscopy screening, detailed contingencies for suspected or newly diagnosed patients as a result of screening were commonly lacking. Personal protective equipment (PPE) recommendations for “front desk” staff and patients in waiting areas was also neglected in some recommendations.

**Table 2:** Recommendations for patient selection for endoscopy during the COVID-19 outbreak.

| Articles grouped by country: | China [4-6] | US [7,8] | UK [9,10] | Spain [11] | Singapore |
| --- | --- | --- | --- | --- | --- |
| **Patient selection in endoscopy** | <u>Triaging:</u><br>- Suspend elective cases and reduce active endoscopy rooms. Urgent or emergency cases only. Postpone all procedures in COVID-19 patients if unnecessary.<br>- Postpone procedures for abdominal pain, vomiting, bloating, diarrhea, coffee ground vomiting or mild PR bleeding, any mild other conditions. Proceed if (i) ingestion of foreign bodies: e.g. batteries, sharp or toxic foreign bodies, (ii) gastrointestinal obstruction caused by foreign bodies, and (iii) endoscopic diagnosis and treatment of major gastrointestinal bleeding. For any other indication e.g. suspected cancers, endoscopist discretion is advised.<br><u>Screening Protocol:</u><br>- Screen all patients for fever at the "front desk". Refer to fever clinic and provide patient with face mask if febrile; axillary body temperature $\geq 37.3^{\circ}\text{C}$ or ear temperature $\geq 37.5^{\circ}\text{C}$. CT Lung if suspicious +/- throat swab.[4] If afebrile, continue risk assessment.<br>- If afebrile, screen for other COVID-19 symptoms, recent travel and close contact history. If suspected COVID-19, perform CT Lung [4]<br><u>PPE recommendation (general staff):</u><br>- Desk staff to wear surgical face mask, caps, impermeable clothing.<br><u>Contingency plan for high-risk patients detected in endoscopy:</u><br>- All patients found to COVID-19 positive to be quarantined in isolation ward. | <u>Triaging:</u><br>- Strongly consider postponing non-urgent or elective cases.<br>- Triage suspected or confirmed COVID-19 patient to designated area. Carers and relatives prohibited from endoscopy department unless necessary.<br><u>Screening Protocol for [6]:</u><br>- 4 questions asked before endoscopy:<br>(i) Fever ($> 37.5^{\circ}\text{C}$) in last 14 days,?<br>(ii) Cough/soar throat/respiratory problems?<br>(iii) Close contact with suspected or confirmed COVID-19 individual? (including family's exposure)<br>(iv) High risk area?<br>- Check body temperature before entering endoscopy.<br>- Classify risk:<br>(i) Low = No symptoms, no contact risks, not from high risk area<br>(ii) Intermediate = One of any positive<br>(iii) High risk = Symptomatic with either contact risk of from high risk area.<br><u>PPE recommendation (general staff):</u><br>- All patients to be offered surgical face masks [6]<br><u>Contingency plan for high-risk patients detected in endoscopy:</u><br>- Not stated. | <u>Triaging:</u><br>- Carry on with all procedures but postpone elective procedures if suspected or confirmed COVID-19.<br>- Hospitals to decide internally about postponing non-urgent procedures.<br><u>Screening protocol [7]:</u><br>(i) Travel history,<br>(ii) Body temperature,<br>(iii) Patients given symptom,<br>(iv) Information sheet and asked to report any symptoms at front desk.<br><u>PPE recommendation (general staff):</u><br>- None stated<br><u>Contingency plan for high-risk patients detected in endoscopy:</u><br>- Not stated. | <u>Triaging:</u><br>- Delay all procedures for 30 days if patients have respiratory symptoms or exposure to contacts regardless of fever unless in emergencies.<br><u>Screening protocol:</u><br>(i) Body temperature,<br>(ii) Respiratory symptoms<br>(iii) High risk contacts<br><u>Contingency plan for high-risk patients detected in endoscopy:</u><br>- Not stated.<br><u>PPE recommendation (general staff):</u><br>- None stated<br><u>Contingency plan for high-risk patients detected in endoscopy:</u><br>- Not stated. | <u>Triaging</u><br>- Non-urgent indications in the following settings to be postponed:<br>1. Patients with acute respiratory Symptoms,<br>2. Exposure in high risk countries<br>3. Suspect COVID-19<br>4. Proven COVID -19<br>- All urgent indications to proceed regardless of COVID-19 status,<br>- Urgency of referral determined by endoscopists.<br><u>Screening protocol:</u><br>(i) Body temperature<br>(ii) Cough<br>(iii) All other COVID-19 symptoms,<br>(iv) Travel history<br>(v) Contact history,<br>- All suspected and confirmed COVID-19 patients to be managed in designated isolation areas.<br><u>PPE recommendation (general staff):</u><br>None stated<br><u>Contingency plan for high-risk patients detected in endoscopy:</u><br>- Not stated. |

### Peri- and Intra-procedural practice recommendations

Stringency of PPE recommendations varied significantly but tended to be more stringent in Asian countries that were previously exposed to the SARS outbreak. The most apparent difference was in the recommendation for respiratory PPEs (Table 2). N95 masks and powered air-purifying respirators (PAPR) were routinely recommended for all endoscopy procedures whereas N95, FFP2 and FFP3 masks were routinely reserved for high-risk patients in US and UK. Also, both US and UK guidelines classified lower endoscopy as low-risk procedures; ASGE recommendations [8] downgrade patients with intermediate risk of COVID-19 to low risk. In these situations, HCWs are permitted to wear surgical face masks in patients with intermediate risk in the US [8], and those with high risk in the UK.[10] Negative pressure ventilation rooms are recommended in some guidelines but not all.

### Post-procedural recommendations

Recommendations for decontamination practices were fairly consistent (Table 3) but some guidelines were more detailed and included air purification measures. Chlorine-containing disinfectants of varying strengths and double-bagging of waste were commonly recommended. General consensus was that standard scope decontamination procedures were adequate. Management of patients post-sedation and recommended PPE for transfer staff were often not mentioned although would be helpful in future revisions.

**Table 3:**
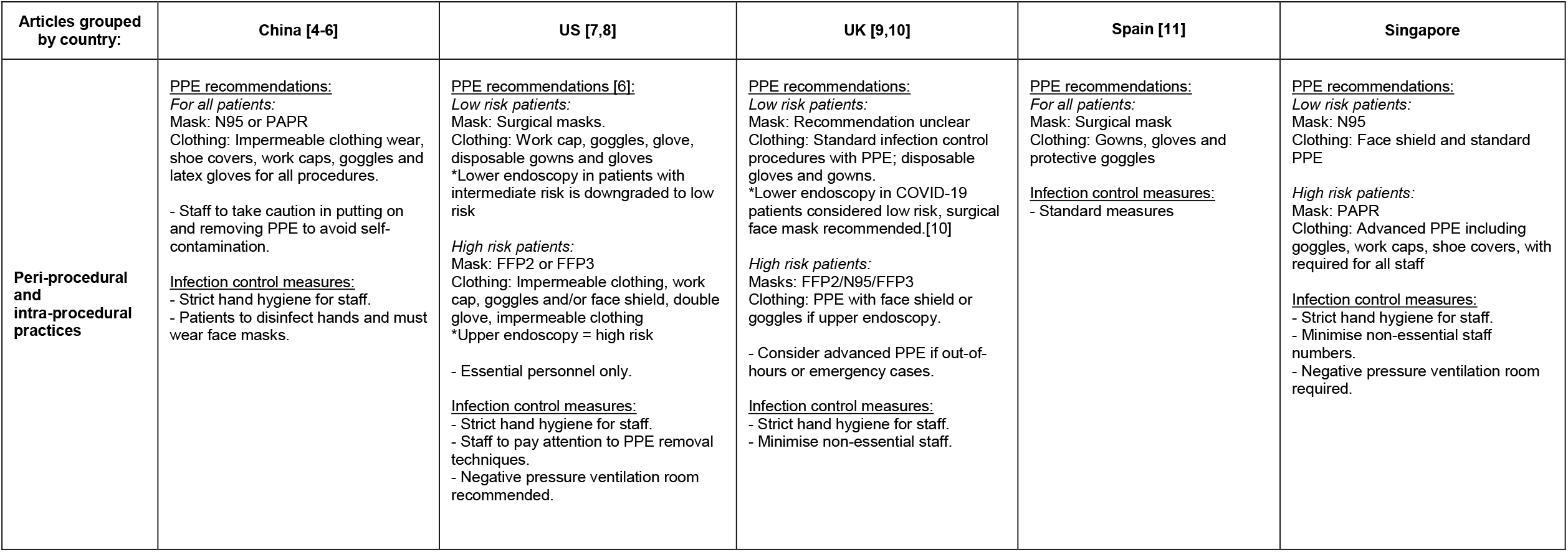
Peri- and intra-procedural recommendations for endoscopy during the COVID-19 outbreak. FFP = Filtering Face Piece

| Articles grouped by country: | China [4-6] | US [7,8] | UK [9,10] | Spain [11] | Singapore |
| --- | --- | --- | --- | --- | --- |
| **Peri-procedural and intra-procedural practices** | <u>PPE recommendations:</u><br>*For all patients:*<br>Mask: N95 or PAPR<br>Clothing: Impermeable clothing wear, shoe covers, work caps, goggles and latex gloves for all procedures.<br>- Staff to take caution in putting on and removing PPE to avoid self-contamination.<br><u>Infection control measures:</u><br>- Strict hand hygiene for staff.<br>- Patients to disinfect hands and must wear face masks. | <u>PPE recommendations [6]:</u><br>*Low risk patients:*<br>Mask: Surgical masks.<br>Clothing: Work cap, goggles, glove, disposable gowns and gloves<br>*Lower endoscopy in patients with intermediate risk is downgraded to low risk<br>*High risk patients:*<br>Mask: FFP2 or FFP3<br>Clothing: Impermeable clothing, work cap, goggles and/or face shield, double glove, impermeable clothing<br>*Upper endoscopy = high risk<br>- Essential personnel only.<br><u>Infection control measures:</u><br>- Strict hand hygiene for staff.<br>- Staff to pay attention to PPE removal techniques.<br>- Negative pressure ventilation room recommended. | <u>PPE recommendations:</u><br>*Low risk patients:*<br>Mask: Recommendation unclear<br>Clothing: Standard infection control procedures with PPE; disposable gloves and gowns.<br>*Lower endoscopy in COVID-19 patients considered low risk, surgical face mask recommended.[10]<br>*High risk patients:*<br>Masks: FFP2/N95/FFP3<br>Clothing: PPE with face shield or goggles if upper endoscopy.<br>- Consider advanced PPE if out-of-hours or emergency cases.<br><u>Infection control measures:</u><br>- Strict hand hygiene for staff.<br>- Minimise non-essential staff. | <u>PPE recommendations:</u><br>*For all patients:*<br>Mask: Surgical mask<br>Clothing: Gowns, gloves and protective goggles<br><u>Infection control measures:</u><br>- Standard measures | <u>PPE recommendations:</u><br>*Low risk patients:*<br>Mask: N95<br>Clothing: Face shield and standard PPE<br>*High risk patients:*<br>Mask: PAPR<br>Clothing: Advanced PPE including goggles, work caps, shoe covers, with required for all staff<br><u>Infection control measures:</u><br>- Strict hand hygiene for staff.<br>- Minimise non-essential staff numbers.<br>- Negative pressure ventilation room required. |

**Table 4:**
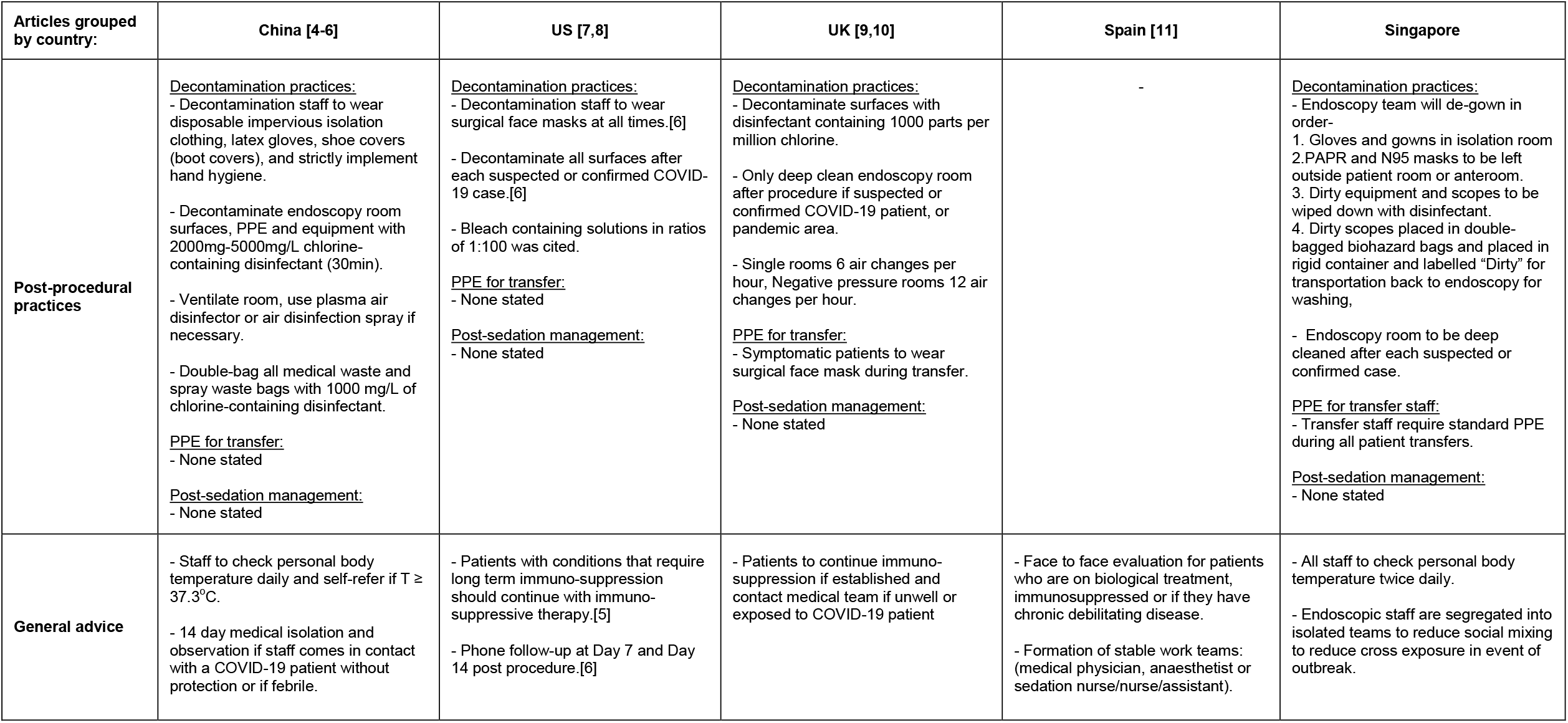
Post-procedural recommendations and general advice for endoscopy during the COVID-19 outbreak.

### General Advice

Most guidelines have commented on, and recommend, the continuation of immune-suppressive medication including biologics in patients already established them. In the event patients become unwell whilst on these medication, the general consensus is for them to seek medical advice urgently. There were no statements from gastrointestinal-related societies against the use of ibuprofen in COVID-19 at the point of writing. 2 of 5 guidelines advised HCWs on how to monitor for signs of self-infection and when to self-report. Only 1 guideline advocated patient follow-up in the community (via telephone) post-procedure.[8]

## DISCUSSION

This review highlights the paucity and need for high-quality evidence. There was little evidence to inform which preventative measures worked best at reducing the incidence of COVID-19 cases in gastrointestinal or endoscopy departments. We have found that current practice is being guided mainly by level 4 and level 5 evidence. Further research in these areas are urgently needed.

Transmission of SARS-CoV2 is through droplets or fomites. It is postulated SARS-CoV2 binds to Angiotensin-Converting Enzyme 2 receptors, and with the assistance of Transmembrane Serine Protease 2, enters cells.[15,16] The virus then replicates in the host and can be detectable in respiratory secretions, stool, blood, tears and urine.[17] In patients with high viral loads, extensive environmental (surfaces, PPE, extractor fans etc.) contamination with viral ribonucleic acid (RNA) has been reported.[2] That said, the most logical and important step to limit the nosocomial transmission of SARS-CoV2 in endoscopy is the screening of patients referred to endoscopy. The early detection of infected patients allows the postponement of procedures until resolution of the infection is achieved, significantly reducing the risk of viral transmission to patients and staff.

However, the median incubation time of the virus is 5.1 days but can extend to more than 11 days (11.5 days = 97.5% percentile), and in the meanwhile they remain asymptomatic but infectious.[18,19] This poses significant problems for screening tools that are heavily dependent on symptomatology. Furthermore, COVID-19 related diarrhoea could also be mistaken for a flare of inflammatory bowel disease or bowel preparation and vice versa. As the spread of COVID-19 becomes more rampant in local communities, screening for travel history may also be limited. Contact screening for exposure to individuals who have symptoms of COVID-19 may prove to be more useful. Nonetheless, this review has not identified any data on the accuracy of question-based screening tools including performance statistics such as area under receiver operating characteristic, positive and negative predictive values, etc.

Given the limitations of question-based screening methods, patient follow-up post-procedure becomes extremely important at detecting “false negatives” that slipped through current processes. Identification of any infected patient post-procedure who was within the window of the viral incubation at the time of endoscopy would have significant implications; undetected transmission to HCWs and other patients in the department must then be investigated. A robust contact screening program is then necessary to contain the spread of COVID-19 among exposed staff and patient contacts. Only 1 guideline identified in this review advised on post-procedure follow-up at Day 7 and Day 14 by telephone. We believe this should be a common feature in all future guidance on COVID-19. As screening methods improve and detection kits become more readily available, biological and radiological screening methods that are advocated by Chinese guidelines may become more efficient at disease detection although would be costly. If COVID-19 becomes a protracted pandemic, one possible solution to help restore normal work flow in endoscopy could be serial screening e.g. 2 throat swabs for viral RNA 2 weeks apart before listing for endoscopy.

The oro-faecal transmission of SARS-CoV2 remains debatable although the virus has been isolated in gastric, duodenal and rectal biopsies with viral RNA detectable in half of all COVID-19 patients.[20] Interestingly, in those that have detectable SARS-CoV2 RNA detectable in their stool, half have diarrhoea and half have normal stool, suggesting a poor correlation between abdominal symptoms and viral RNA positivity.[3] Viable viral culture from stool samples are also lacking [3], and we have found no evidence of the transmission of either SARS-CoV or SARS-CoV2 through endoscopy. However, such reports may surface in the future as infection becomes more common. Nonetheless, the index of suspicion for oro-faecal transmission remains high and this is particularly relevant for lower endoscopy. Both US and UK guidelines regard lower gastrointestinal endoscopy as low-risk procedures and therefore are less stringent with respiratory PPEs; in the UK lower endoscopy are not regarded as aerosol generating procedures. Chinese and Singaporean guidelines may have erred on the side of caution and advocated the use of FFP2/N95/PAPR masks because microbial contamination of air and PPE after lower endoscopy has been reported [21,22]. One study reported that applying suction during the removal of biopsy forceps decreased environmental bacterial contamination.[21] In our experience, resource allocation for staff education, time for decontamination, and management of the physical and mental wellbeing of HCWs were also important in maintaining endoscopy services and should not be underestimated.

This systematic review has a several limitations. At the time of this search, any advice communicated by societies or government bodies through email or circulars, may not have been identified in the searches. Also, differences in health policy, resource availability and health economics may have contributed to the heterogeneity in guidelines between countries.

In conclusion, stronger evidence to inform current practice and robust guidelines are urgently needed to prevent the transmission of SARS-CoV2 in gastrointestinal endoscopy departments worldwide.

## Data Availability

All data has been provided or referenced in the manuscript.

## Acknowledgements

JO is supported by the W.D. Armstrong Doctoral Fellowship from Cambridge University and a development grant from the National University of Singapore. The views expressed are those of the author(s) and not necessarily those of the NHS or the Department of Health.

---

**Supplementary Table 1:** List of gastroenterology and endoscopy societies searched for guidelines or advice on endoscopy in a COVID-19 outbreak. * denotes societies that have published guidance on COVID-19.

| No. | COVID-19 cases<br>(as of 16 Mar 20) | Country | Gastroenterology and endoscopy<br>societies searched (*n* = 22) |
| --- | --- | --- | --- |
| 1 | 81,020 | China * | Chinese Society of Gastroenterology,<br>Chinese Medical Association |
| 2 | 23,980 | Italy | Società Italiana di Gastroenterologia,<br>Società Italiana Endoscopia Digestiva |
| 3 | 13,938 | Iran | Iranian Association for Gastroenterology<br>and Hepatology |
| 4 | 8,236 | South Korea | Korean Society of Gastroenterology,<br>Korean Society of Gastrointestinal<br>Endoscopy |
| 5 | 7,753 | Spain * | Sociedad Española de Patología<br>Digestiva |
| 6 | 5,423 | France | Société Nationale Française de<br>Gastroenterologie |
| 7 | 4,838 | Germany | Deutsche Gesellschaft für<br>Gastroenterologie, Verdauungs- und<br>Stoffwechselkrankheiten |
| 8 | 3,774 | USA * | American Gastroenterological<br>Association, American College of<br>Gastroenterology, American Society of<br>Gastrointestinal Endoscopy |
| 9 | 2,200 | Switzerland | Schweizerische Gesellschaft für<br>Gastroenterologie SSG Société Suisse<br>de Gastro-entérologie |
| 10 | 1,391 | UK * | British Society of Gastroenterology,<br>Public Health England |
| 11 | 1,135 | Netherlands | Nederlandse Vereniging voor Gastro-<br>enterologie |
| 12 | 1,077 | Norway | Norsk Gastroenterologisk Forening |
| 13 | 1,032 | Sweden | Svensk Gastroenterologisk Förening |
| 14 | 886 | Belgium | Belgian Society of Gastrointestinal<br>Endoscopy, Société Royale Belge de<br>Gastro-Entérologie |
| 15 | 875 | Denmark | Dansk Selskab for Gastroenterologi og<br>Hepatologi |
| **Other societies searched (*n* = 6) :**<br>Asian Pacific Association of Gastroenterology<br>European Society of Gastrointestinal Endoscopy<br>Scandinavian Association for Digestive Endoscopy<br>United European Gastroenterology<br>World Gastroenterology Organisation<br>World Endoscopy Organisation |  |  |  |

PRISMA 2009 Checklist
| Section/topic | # | Checklist item | Reported on page # |
| --- | --- | --- | --- |
| **TITLE** |  |  |  |
| Title | 1 | Identify the report as a systematic review, meta-analysis, or both. | 1 |
| **ABSTRACT** |  |  |  |
| Structured summary | 2 | Provide a structured summary including, as applicable: background; objectives; data sources; study eligibility criteria, participants, and interventions; study appraisal and synthesis methods; results; limitations; conclusions and implications of key findings; systematic review registration number. | 2 |
| **INTRODUCTION** |  |  |  |
| Rationale | 3 | Describe the rationale for the review in the context of what is already known. | 3 |
| Objectives | 4 | Provide an explicit statement of questions being addressed with reference to participants, interventions, comparisons, outcomes, and study design (PICOS). | 3 |
| **METHODS** |  |  |  |
| Protocol and registration | 5 | Indicate if a review protocol exists, if and where it can be accessed (e.g., Web address), and, if available, provide registration information including registration number. | Not applicable |
| Eligibility criteria | 6 | Specify study characteristics (e.g., PICOS, length of follow-up) and report characteristics (e.g., years considered, language, publication status) used as criteria for eligibility, giving rationale. | 5 |
| Information sources | 7 | Describe all information sources (e.g., databases with dates of coverage, contact with study authors to identify additional studies) in the search and date last searched. | 4, 5, 6 |
| Search | 8 | Present full electronic search strategy for at least one database, including any limits used, such that it could be repeated. | 4 |
| Study selection | 9 | State the process for selecting studies (i.e., screening, eligibility, included in systematic review, and, if applicable, included in the meta-analysis). | 4, 5 |
| Data collection process | 10 | Describe method of data extraction from reports (e.g., piloted forms, independently, in duplicate) and any processes for obtaining and confirming data from investigators. | 5 |
| Data items | 11 | List and define all variables for which data were sought (e.g., PICOS, funding sources) and any assumptions and simplifications made. | 4, 5 |
| Risk of bias in individual studies | 12 | Describe methods used for assessing risk of bias of individual studies (including specification of whether this was done at the study or outcome level), and how this information is to be used in any data synthesis. | 5 |
| Summary measures | 13 | State the principal summary measures (e.g., risk ratio, difference in means). | 4, 5 |
| Synthesis of results | 14 | Describe the methods of handling data and combining results of studies, if done, including measures of consistency (e.g., $I^2$) for each meta-analysis. | Meta-analyses not done |

PRISMA 2009 Checklist
| Section/topic | # | Checklist item | Reported on page # |
| --- | --- | --- | --- |
| Risk of bias across studies | 15 | Specify any assessment of risk of bias that may affect the cumulative evidence (e.g., publication bias, selective reporting within studies). | 5 |
| Additional analyses | 16 | Describe methods of additional analyses (e.g., sensitivity or subgroup analyses, meta-regression), if done, indicating which were pre-specified. | Meta-analyses not done |
| **RESULTS** |  |  |  |
| Study selection | 17 | Give numbers of studies screened, assessed for eligibility, and included in the review, with reasons for exclusions at each stage, ideally with a flow diagram. | 6 |
| Study characteristics | 18 | For each study, present characteristics for which data were extracted (e.g., study size, PICOS, follow-up period) and provide the citations. | 6 |
| Risk of bias within studies | 19 | Present data on risk of bias of each study and, if available, any outcome level assessment (see item 12). | Not applicable |
| Results of individual studies | 20 | For all outcomes considered (benefits or harms), present, for each study: (a) simple summary data for each intervention group (b) effect estimates and confidence intervals, ideally with a forest plot. | 6, Table 1 |
| Synthesis of results | 21 | Present results of each meta-analysis done, including confidence intervals and measures of consistency. | Not applicable |
| Risk of bias across studies | 22 | Present results of any assessment of risk of bias across studies (see Item 15). | Not applicable |
| Additional analysis | 23 | Give results of additional analyses, if done (e.g., sensitivity or subgroup analyses, meta-regression [see Item 16]). | 6, 7, 8 |
| **DISCUSSION** |  |  |  |
| Summary of evidence | 24 | Summarize the main findings including the strength of evidence for each main outcome; consider their relevance to key groups (e.g., healthcare providers, users, and policy makers). | 8, 9, 10, 11 |
| Limitations | 25 | Discuss limitations at study and outcome level (e.g., risk of bias), and at review-level (e.g., incomplete retrieval of identified research, reporting bias). | 11 |
| Conclusions | 26 | Provide a general interpretation of the results in the context of other evidence, and implications for future research. | 8, 9, 10, 11 |
| **FUNDING** |  |  |  |
| Funding | 27 | Describe sources of funding for the systematic review and other support (e.g., supply of data); role of funders for the systematic review. | 12 |

From: Moher D, Liberati A, Tetzlaff J, Altman DG, The PRISMA Group (2009). Preferred Reporting Items for Systematic Reviews and Meta-Analyses: The PRISMA Statement. PLoS Med 6(7): e1000097. doi:10.1371/journal.pmed1000097
For more information, visit: www.prisma-statement.org.

